# An immuno-enrichment free, validated quantification of tau protein in human CSF by LC-MS/MS

**DOI:** 10.1101/2021.05.10.21252513

**Authors:** Wade Self, Khader Awwad, John Paul Savaryn, Michael Schulz

## Abstract

**Aims:** Tau protein is a key target of interest in developing therapeutics for neurodegenerative diseases. Here, we sought to develop a method that quantifies extracellular tau protein concentrations human cerebrospinal fluid (CSF) without antibody-based enrichment strategies.

**Results:** We demonstrate that the fit-for-purpose validated method in Alzheimer’s Disease CSF is limited to quasi quantitative measures of tau surrogate peptides. We also provide evidence that CSF total Tau measures by LC-MS are feasible in the presence of monoclonal therapeutic antibodies in human CSF.

**Conclusion:** Our Tau LC-MS/MS method is a translational bioanalytical tool for assaying target engagement and pharmacodynamics for anti-tau antibody drug development campaigns.

## Introduction

No effective disease-modifying treatments exist for progressive neurodegenerative diseases associated with aging. A unifying pathological observation in these disease states is the accumulation of misfolded insoluble protein aggregates within brain regions that degenerate, leading to neurological symptoms. In a group of disorders termed “tauopathies” that includes Alzheimer’s Disease (AD), these protein aggregates are characterized by the presence of tau protein [1]. The presence of tau aggregation strongly correlates with neuronal atrophy in specific brain regions and cognitive decline in AD [2,3], suggesting that tau is a key molecular driver of disease progression in tauopathies. In AD, the presence of tau pathology follows a specific pattern, where tau aggregates are first observed in the entorhinal cortex, followed by “spread” to the hippocampus and later to the cortex [4]. A leading hypothesis to explain this phenomenon is that pathological tau protein is released extracellularly and spreads via anatomically connected neuronal pathways in a prion-like mechanism [5,6]. These observations suggest that targeting extracellular, pathogenic tau species may be a viable therapeutic strategy for disease modification in tauopathies. Although the exact tau species to target remains elusive, multiple monoclonal antibodies to bind and clear extracellular tau species are in clinical development [7,8]. These campaigns utilize a “pan-tau” approach, with antibody epitopes that bind the N-terminus of tau protein.

To understand the efficacy of a pan-tau targeting approach with monoclonal antibodies, direct assessments of pharmacodynamics are crucial to incorporate into human clinical trials. In humans, tau protein is present in the cerebrospinal fluid (CSF) and provides a surrogate measure for tau activity within the brain [9], presenting an opportunity to utilize an accessible biofluid to develop pharmacodynamic assays. However, tau protein is a complex molecule that makes generation of “pan-tau” measurements challenging. Tau consists of six isoforms in the brain due to alternative splicing of the *MAPT* gene, and further post-translational modifications give rise to a complex pool of Tau proteoforms in the central nervous system [10-12]. Further, proteolytic cleavage of tau results in multiple tau protein fragments that current data suggest are in low abundance in CSF on the order of single picograms to nanograms per milliliter [13-15]. To overcome these analytical challenges, multiple ligand-binding assays have been developed with sufficient sensitivity for CSF tau bioanalysis [14,16]. Despite their sensitivity, multiple limitations exist in using tau LBA assays as a pharmacodynamic assay, as interpretation of results are limited to a single tau fragment that contains the capture and detection antibody epitope and may not fully represent a “pan-tau” molecular signature. Further, the epitope of the therapeutic antibody must be considered when designing an appropriate tau LBA, as therapeutic antibody binding to tau may interfere with the binding of CSF tau to capture and detection antibodies.

One solution to overcome these limitations is to develop a multiplexed assay that captures information across the entire tau amino acid sequence to fully analyze differential tau fragment pharmacodynamic responses to experimental treatment antibodies using tandem liquid chromatography-mass spectrometry (LC-MS)-based methods. Indeed, some have established immunoenrichment-free LC-MS methods to quantify multiple CSF tau surrogate peptides using a partial perchloric acid (PCA) precipitation combined with intact protein solid phase extraction (SPE) for sample preparation[17,18]. Using these methods as a foundation, we aimed to increase the throughput and robustness of the method through alterations in sample preparation and the LC-MS analysis to enable clinical applicability. We were particularly interested in optimizing conditions to identify surrogate peptides that correspond to the N-Terminus of Tau, as these are where the epitopes for most “Pan-Tau” targeting monoclonal antibodies are located. In this report, we determined the fit-for-purpose biomarker assay capabilities of this LC-MS assay in AD CSF using a standard curve of recombinant, intact 2N4R tau in artificial CSF matrix, including what is, to our knowledge, a first evaluation of parallelism for LC-MS tau quantification. Moreover, we tested the capability of our LC-MS assay to measure CSF Tau in the presence of therapeutic tau monoclonal antibody to mimic samples available for pharmacodynamic analysis in human clinical trials.

## Methods

### Recombinant Protein Standards

Tau 441 recombinant protein was produced at the AbbVie Biotherapeutics Center (Worchester, MA), in both an unlabeled and N15 stable-isotope labeled form, per previously-established protocols[19]. This tau protein standard corresponds to the 441 amino acid 2N4R isoform of human tau protein.

### Description of Biofluid Samples

Samples of pooled CSF from non-diseased individuals and human serum samples were commercially purchased from BioIVT. Deidentified human cerebrospinal fluid samples from persons with Alzheimer’s Disease of various ages (range: 60-77), gender (male and female), and ethnicity (Caucasian, African American, Hispanic) were commercially purchased from Precision Med Inc., with total CSF Tau concentrations measured by the V-Plex Human Total Tau MSD Kit (Mesoscale Diagnostics) ranging from 368 - 1265 pg/mL. All samples were stored at -80 □C before use.

### Pre-Validation Optimization Studies

#### PCA Precipitation Optimization

Two serial dilutions of Tau 441 recombinant Protein in an artificial CSF matrix consisting of 0.5% human serum in phosphate-buffered saline (PBS) was performed. Samples were processed identically to the Barthelemy *et al*. sample preparation method [17] (Figure 1A) with differences in the amount of PCA used for protein precipitation in each serial dilution series (2.5% vs. 1% PCA final volume).

**Figure 1.**
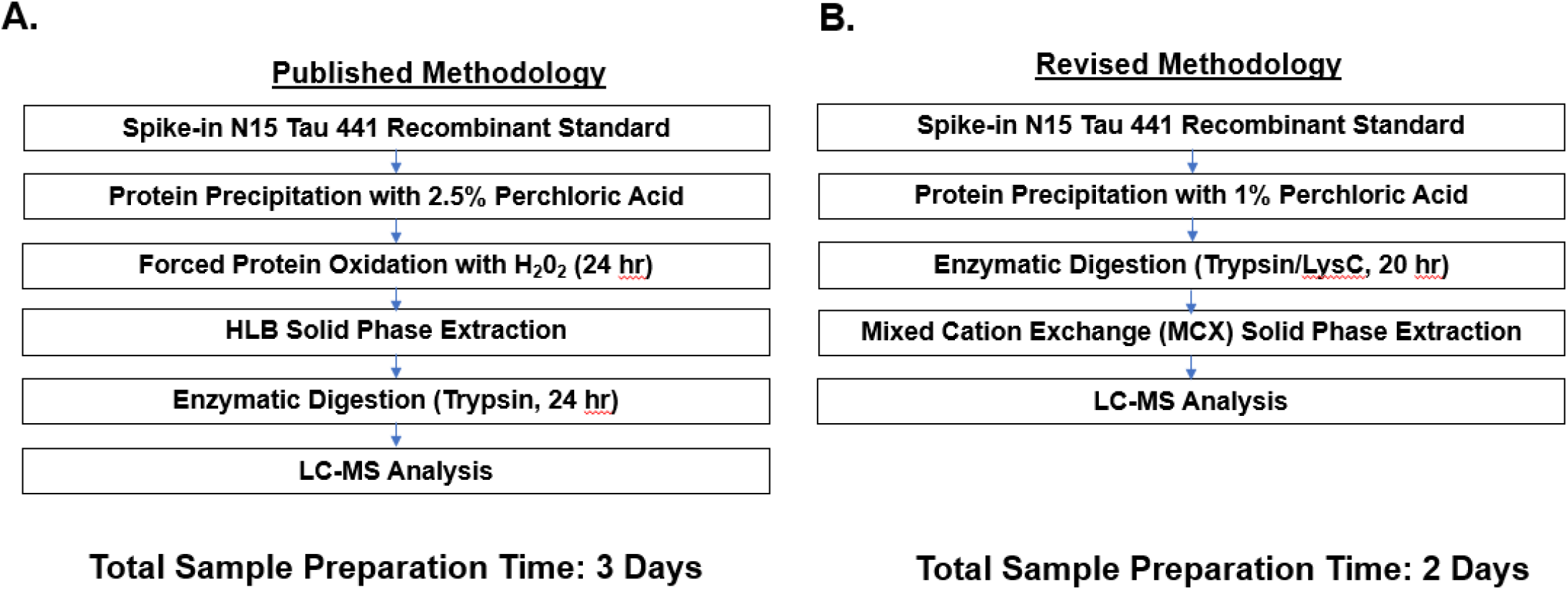
Sample preparation workflow for downstream CSF Total Tau LC-MS analysis. A. Protocol published in the literature that served as a foundation for the final analysis. B. Adaptations from the published protocol that were used as the final assay protocol for validation.

#### LC-MS/MS Analysis

20 µL of sample was loaded onto a PepMap 300 C18 HPLC Column (300 um x 5 cm, 5 um, Thermo) using an Ultimate 3000 UHPLC autosampler at a flow rate of 5 ul/min. Mobile phase consisted of 0.1% Formic Acid in water (Solvent A) and 0.1% Formic Acid in acetonitrile (Solvent B). Analyte was loaded on stationary phase for 3 minutes with 2% B, followed by a mobile phase gradient from 2-40% B over 70 minutes to elute surrogate peptides from a C18 stationary phase EASY-spray analytical column (75 um ID, 15 cm, Thermo) at a flow rate of 0.3 ul/min at 45 □C. The analytical column was washed with 70% B for five minutes and re-equilibrated with 2% B for 15 minutes.

Mass spectrometry analysis was performed on a Q Exactive Quadrupole Orbitrap instrument (Thermo) operating in positive electrospray ionization mode. A parallel reaction monitoring (PRM) method with 17,500 resolution (*m/z* = 200), 0.7 *m/z* mass tolerance, automatic gain control of 2e5, and HCD fragmentation was used to analyze 5 surrogate tau peptides and their co-eluting N15-labeled internal standards with an error of < 5 ppm (Table 1).

**Table 1.**
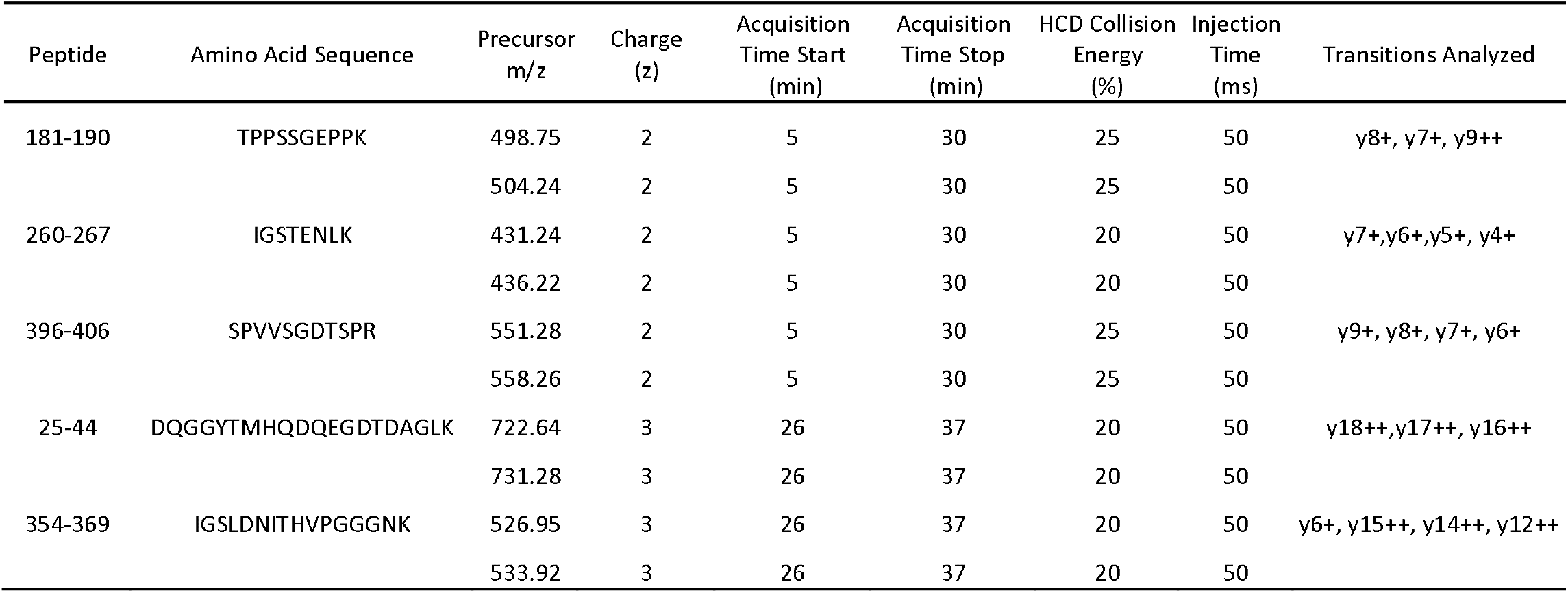
Mass Spectrometry parameters on the Thermo Q Exactive for Pre-Validation experiments using nanoLC-MS/MS.

### CSF Total Tau Bioanalysis Method

The sample preparation workflow was optimized from multiple reports in the literature[17,20], with modifications (Figure 1). These include: 1.) Changing the final concentration of perchloric acid (PCA) in CSF samples to 1%, 2.) Eliminating the intact protein solid phase extraction (SPE) and forced oxidation steps, 3.) Performing enzymatic digestion with Trypsin/LysC, and 4.) Adding a peptide-level SPE using mixed cation exchange (MCX) chromatography.

#### Calibration and Quality Control Standards

A reference standard curve was prepared for each analytical run by performing a serial dilution of Tau 441 Recombinant Protein in an artificial CSF matrix consisting of 0.5% human serum in phosphate-buffered saline (PBS). Quality Control (QC) samples were prepared by spiking-in Tau 441 Recombinant Protein into artificial CSF at low (1,000 pg/mL), medium (4,000 pg/mL), and high (16,000 pg/mL) concentrations. Reference standard curve and QC samples were run in duplicate.

#### Sample Preparation

50 µL of N15-labeled Tau internal standard solution was added to 500 µL of human CSF, reference standard, and QC samples. The final mass of N15-labeled Tau in CSF samples was 5 ng (concentration: 10 ng/mL). Samples were treated with 10% perchloric acid (Millipore Sigma, St. Louis, MO) to a final concentration of 1% per sample, and incubated at 4□ C for 15 minutes with shaking at 1000 rpm using a Thermomixer (Eppendorf). Samples were then placed in a tabletop centrifuge and spun at 16,000 x g for 15 minutes. The sample supernatants, containing tau protein, were then transferred to a Lo-bind 96 well plate (Eppendorf). 275 µL of 1 M Tris HCl (Millipore Sigma, St. Louis, MO) was added to each sample to bring the solution to a pH = 8. 10 µg of Sequencing Grade Trypsin/LysC (Promega) was added to each sample and incubated 20 hours at 37□ C, with shaking at 1000 rpm in a Thermomixer. Samples were then cooled to room temperature and the digestion reaction was quenched using 125 µL of 20% phosphoric acid (Sigma). Samples were then transferred to individual wells of a Mixed Cation Exchange (MCX) SPE plate (Waters, Milford, MA) and washed per the manufacter’s protocol. Samples were eluted from MCX columns using 95% Methanol/5% ammonium hydroxide into a new Lo-bind 96 well plate, dried under vacuum at 45 □C (LabConco), and resuspended in 80 µL of 2% Acetonitrile/0.1% Triflouroacetic Acid (TFA) for LC-MS/MS analysis.

#### LC-MS/MS Analysis

30 µL of sample was loaded onto an HSS T3 stationary phase column (2.1 mm x 15 cm, 1.7 µm, Waters Inc.) using a Vanquish UHPLC System (Thermo) at a flow rate of 200 µl/minute at 60 □C. Mobile phase consisted of 0.1% Formic Acid in water (Solvent A) and 0.1% Formic Acid in acetonitrile (Solvent B). Analyte was loaded on stationary phase for 2 minutes with 2% B, followed by a mobile phase gradient from 2-45% B over 11 minutes to elute surrogate peptides. The analytical column was washed with 95% B for two minutes and re-equilibrated with 2% B for 2 minutes.

Mass spectrometry analysis was performed on a Fusion Orbitrap Tribrid instrument (Thermo) operating in positive electrospray ionization mode (Ion Spray Voltage: 3700 V, Capillary Temperature 320 □C, Sheath Gas: 35 PSI, Auxillary Gas: 8 PSI, Radio Lens RF: 40). A parallel reaction monitoring (PRM) method with 30,000 resolution (*m/z* = 200), 0.7 *m/z* mass tolerance, automatic gain control of 5e5, and HCD fragmentation was used to analyze the 7 surrogate tau peptides and their co-eluting N15-labeled internal standards with an error of < 5 ppm (Table 2).

**Table 2.**
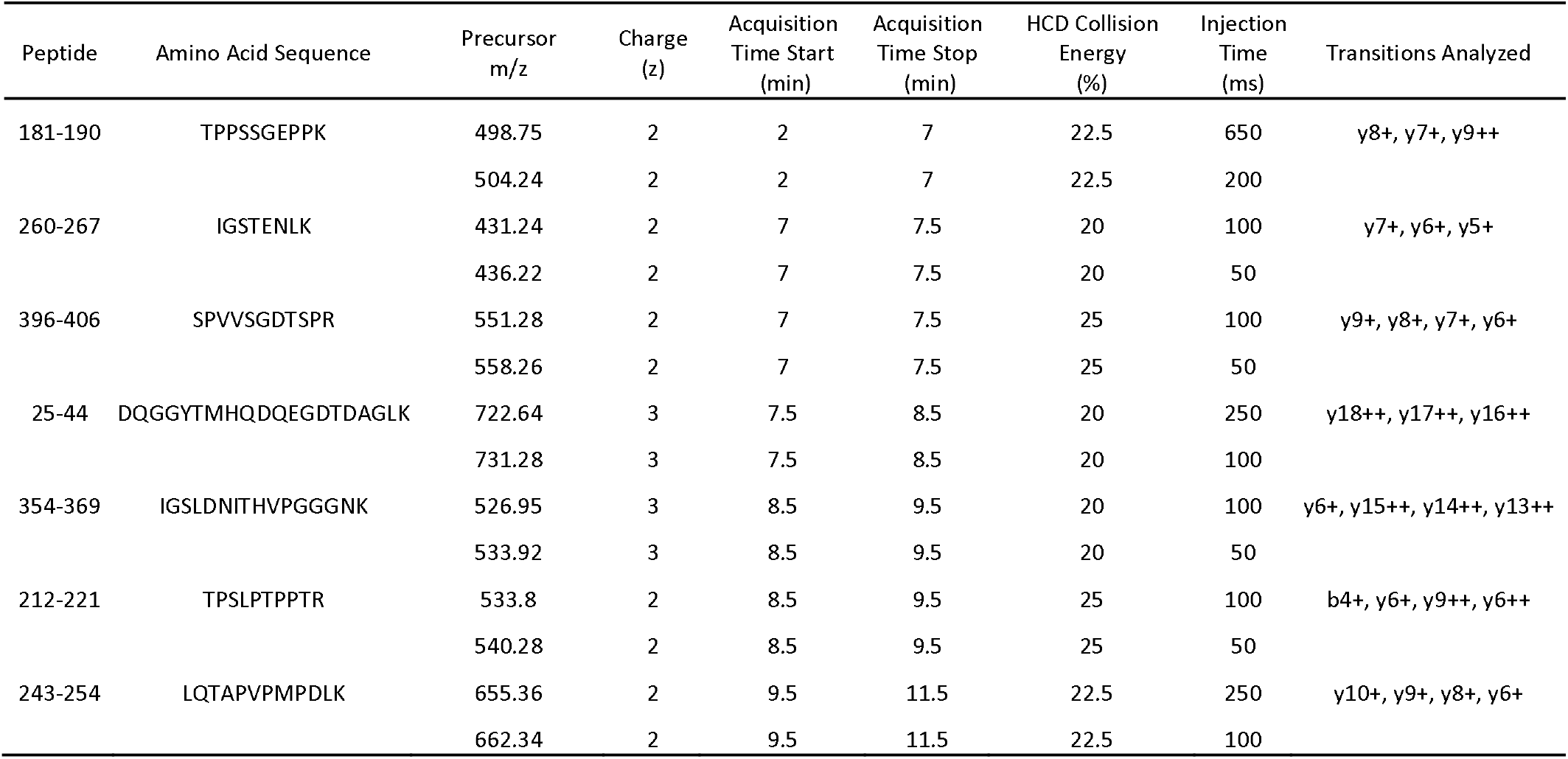
Mass Spectrometry parameters on the Thermo Fusion Tribrid for final Tau LC-MS/MS method validation.

### Data Analysis

Data was collected using XCalibur software (Thermo), and chromatograms were extracted using Skyline software (Seattle, WA). The top 3-4 transitions for each precursor ion were analyzed. All endogenous tau peptide signals (light) were normalized to internal standard (heavy) and imported into Microsoft Excel for data analysis. The calculated concentration for each sample was determined by linear regression of the light:heavy peptide ratio to Tau 441 recombinant protein concentration in the reference standard curve. Coefficient of variation (CV) is reported for sample preparation replicates, and mean bias was calculated by determining the relative error of the calculated concentration to the known peptide concentration for each sample.

### Method Validation

#### Analytical Run Acceptance Criteria

An analytical run was accepted if the following criteria were met: 1.) A coefficient of determination from the reference standard curve of ≥ 0.95, 2.) QC High and Medium samples showed a coefficient of variance (CV) and mean bias (absolute value of % relative error) ≤ 20%, and 3.) QC Low samples showed CV and mean bias levels ≤ 30%.

#### Accuracy and Precision in Human CSF with recombinant tau standard

The precision of CSF tau peptide measurements and accuracy of calculated concentrations against the reference standard curve were determined in non-disease, pooled human CSF samples. Four conditions were assayed in an analytical run: endogenous protein concentration and three concentrations of recombinant tau 441 spiked into CSF samples: 1,000 pg/mL, 4,000 pg/mL, and 16,000 pg/mL. Samples were prepared in triplicate, and the experiment was repeated across three independent analytical runs. The calculated concentration for each sample is reported as the mean of all three analytical runs, and the variance was calculated across all sample preparation replicates. The accuracy of each spike-in concentration level was determined by calculating the relative error of comparing the known tau 441 recombinant protein spike-in concentration to the difference of the calculated peptide concentration and the endogenous peptide concentration. The predetermined acceptance criteria for accuracy and precision of recombinant tau 441 in CSF matrix was a calculated concentration value with mean bias ≤ 30% and CV ≤ 30%.

#### Parallelism and Dilutional Linearity of endogenous tau protein in Alzheimer’s Disease CSF

Parallelism of endogenous tau peptide signal was tested in two AD CSF samples with high concentrations of Tau (AD1, AD2). AD1 and AD2 were subject to 3 1:2 serial dilutions in a pool of AD CSF from donor samples AD 4, AD8, AD9. This pooled AD CSF had calculated tau peptide concentrations that were 3X lower than AD1 and AD2 peptide concentrations. CSF Tau peptide concentrations for the undiluted and diluted CSF samples were calculated from the reference standard curve. To determine parallelism, the slopes of the reference standard curve samples were compared to the slopes of diluted AD1 and AD2 samples. Samples were considered parallel if the calculated slopes of each curve were within 30% of the slope of the reference standard curve. Linearity was determined for Alzheimer’s Disease CSF by a coefficient of determination from the diluted AD CSF standard curve of ≥ 0.8.

#### Biomarker Assay Categorization

The fit-for-purpose utility of this assay was determined for each of the 7 tau peptides independently by the previously defined biomarker criteria [21]. Each surrogate peptide analysis was treated as an independent biomarker assay when determining fit-for-purpose. This investigation determined if the assay was either a relative quantification, quasi quantitative, or qualitative biomarker assay.

#### Alzheimer’s Disease CSF Biological Variability

11 AD CSF samples were analyzed in sample preparation duplicates across 2 independent analytical runs. The CV of the calculated concentrations for each AD CSF sample is reported for the 4 sample preparation replicates across the 2 analytical runs.

#### Therapeutic Antibody Interference

Non-diseased pooled CSF samples were incubated with ABBV-8E12 [8] at varying concentrations (0 µg/mL, 2.5 µg/mL, 5 µg/mL) for 1 hour at 37 °C with shaking at 800 rpm in a Thermomixer before 1% PCA precipitation. Three sample preparation replicates were analyzed for each condition. Antibody interference was determined by calculating the mean bias of the calculated CSF tau concentrations in 2.5 µg/mL and 5 µg/mL samples compared to the 0 µg/mL. Antibody interference was defined as a mean bias > 30% between the calculated concentrations of samples with ABBV-8E12 compared to 0 µg/mL samples.

## Results

### CSF Total Tau Assay Modifications

Before assay validation, we optimized multiple parameters from the published protocol by Barthelemy *et al*. with the goal of improving sample throughput and operational efficiency (Figure 1A) [17]. Previous reports that used PCA precipitation for tau protein purification suggested that 1% PCA precipitation improved tau recovery compared to 2.5% PCA final concentration [22]. To determine how different PCA concentrations impact assay sensitivity, we compared the lower limit of quantification of recombinant Tau 441 in artificial CSF using 1% PCA and 2.5% PCA [17]. For 3 out of the 5 surrogate peptides assayed across the tau protein sequence (25-44, 181-190, 354-369), we observed a lower LLOQ using 1% PCA compared to 2.5% PCA, while two surrogate peptides (260-267, 396-406) showed no changes in sensitivity (Table 3). 1% PCA concentration conditions also enabled the use of a low volume of 1 M Tris HCl to buffer the tau-containing supernatant to pH = 8. These conditions allowed for a direct digest of buffered solution using Trypsin/Lys-C in a Lo-Bind 96 well plate format, improving standardization of the assay to mimic existing workflows for pharmacokinetics-based bioassays by LC-MS.

**Table 3.** Optimization of Perchloric Acid Precipitation conditions. LLOQ was defined as the sample in reference standard curve (Tau 441 in artificial CSF) where mean bias of the measured concentration value relative to the known Tau 441 concentration in reference standard was ≤ 25%. CV for all samples was ≤ 20%.

| Peptide | 2.5% LLOQ<br>(pg/mL) | Mean Bias<br>(\| % RE \|) | 1% PCA LLOQ<br>(pg/mL) | Mean Bias<br>(\| % RE \|) |
| --- | --- | --- | --- | --- |
| 25-44 | 1400 | 6 | 700 | 3 |
| 181-190 | 700 | 13 | 350 | 20 |
| 260-267 | 350 | 4 | 350 | 25 |
| 354-369 | 700 | 3 | 175 | 8 |
| 396-406 | 350 | 6 | 350 | 17 |

Because two surrogate tau peptides of interest (25-44, 243-254) contained a methionine in the primary amino acid sequence, we analyzed the abundance of unoxidized, mono-oxidized (+16 *m/z*) and di-oxidized (+32 *m/z*) in our workflow with and without forced oxidation by incubation with H_2_0_2_ in the SPE step described in the original Barthelemy et al. protocol [17] (Table 5). We demonstrated the ability of forced oxidation with H_2_0_2_ to shift the predominant peptide species to the +16 *m/z* mono-oxidized species. However, the percent abundance of the +16 *m/z* species to total peptide signal was similar to the unmodified peptide abundance without the forced oxidation step. The relative abundance of each peptide, as measured by LC-MS/MS peak area, remained consistent after re-analyzing these samples after 72 hours of incubation in the autosampler, with < 5% change in relative signal abundance for each quantified peptide. These data suggest no further peptide oxidation occurred within this timeframe (Table 4). Therefore, we concluded that the forced oxidation step is not necessary for improving the abundance of methionine-containing tau surrogate peptides in this workflow. Finally, sample clean-up with mixed cation exchanged SPE at the peptide level resulted in more consistent sample dry-down times compared to SPE at the intact protein step (data not shown). Taken together, these data demonstrate modifications that improve sample preparation consistency and throughput for CSF Tau bioanalysis (Figure 1B).

**Table 4.** Forced Oxidation Optimization, using the protocol established by Barthelemy et al. [17] (Figure 1A). Peak area % was determined by dividing the peak area for a single precursor across the sum of the peak area across all precursor ions analyzed. 722.63, 655.36 m/z = unoxidized methionine. 727.92, 663.36 m/z = monoxidized methionine (+16 Da). 733.30, 671.36 m/z = deoxidized methione (+32 Da).

| (25-44) DQGGYT**M**HQDQEGDTDAGLK |  | 722.63 m/z | 727.92 m/z | 733.30 m/z | 722.63 m/z | 727.92 m/z | 733.30 m/z |
| --- | --- | --- | --- | --- | --- | --- | --- |
| Oxidation Status | Time in autosampler (hr) | Peak Area | Peak Area | Peak Area | Peak Area % | Peak Area % | Peak Area % |
| Forced | 0 | 2.62E+04 | 4.95E+06 | 6.93E+05 | 0.46 | 87.32 | 12.22 |
| Forced | 24 | 6.97E+03 | 8.87E+06 | 5.14E+05 | 0.07 | 94.45 | 5.48 |
| Forced | 96 | 0.00E+00 | 5.11E+06 | 3.42E+05 | 0 | 93.73 | 6.27 |
| None | 0 | 2.22E+07 | 2.31E+06 | 0.00E+00 | 90.56 | 9.44 | 0 |
| None | 24 | 2.67E+07 | 3.00E+06 | 4.80E+03 | 89.90 | 10.08 | 0.02 |
| None | 72 | 1.64E+07 | 2.41E+06 | 4.69E+03 | 87.17 | 12.80 | 0.03 |
| (243-254) LQTAPVP**M**PDLK |  | 655.36 m/z | 663.36 m/z | 671.36 m/z | 655.36 m/z | 663.36 m/z | 671.36 m/z |
|  |  | Peak Area | Peak Area | Peak Area | Peak Area % | Peak Area % | Peak Area % |
| Forced | 0 | 3.17E+03 | 2.51E+07 | 3.01E+06 | 0.01 | 89.30 | 10.69 |
| Forced | 24 | 3.12E+04 | 3.90E+07 | 3.31E+06 | 0.07 | 92.10 | 7.83 |
| Forced | 96 | 0.00E+00 | 1.95E+07 | 1.69E+06 | 0 | 92.01 | 7.99 |
| None | 0 | 4.08E+07 | 6.53E+06 | 7.56E+03 | 86.20 | 13.78 | 0.02 |
| None | 24 | 6.22E+07 | 1.19E+07 | 2.09E+05 | 83.72 | 16.00 | 0.28 |
| None | 72 | 1.21E+07 | 1.63E+06 | 3.14E+04 | 87.89 | 11.88 | 0.23 |

**Table 5.** Tau LC-MS Accuracy and Precision Validation using recombinant tau 441 in non-disease CSF. Reported values represent the inter-assay mean endogenous concentration, mean bias, and variance across the three analytical runs (n = 9 replicates)

| Surrogate Peptide | Functional Domain | <u>Measured Concentration (pg/mL)</u> |  | <u>Measured Concentration Mean Bias (\| % RE \|)</u> |  |  |
| --- | --- | --- | --- | --- | --- | --- |
|  |  | aCSF Calibration Range | Endogenous (% CV) | Endogenous + 1,000 pg/mL | Endogenous + 4,000 pg/mL | Endogenous + 16,000 pg/mL |
| 25-44 | N-Terminus | 1,250 – 40,000 | 7,843<br>(2) | 76 | 4 | 17 |
| 181-190 | Mid-Domain | 312 – 40,000 | 12,358<br>(5) | 127 | 11 | 19 |
| 212-221 | Mid-Domain | 625 – 40,000 | 4,972<br>(9) | 70 | 4 | 12 |
| 243-254 | MTBR | 625 – 40,000 | 457*<br>(23) | 92 | 17 | 18 |
| 260-267 | MTBR | 625 – 40,000 | 1,700<br>(15) | 130 | 11 | 21 |
| 354-369 | MTBR | 625 – 40,000 | 1,740<br>(9) | 107 | 16 | 22 |
| 396-406 | C-Terminus | 625 – 40,000 | 797<br>(68) | 94 | 18 | 21 |

\* Below linear range of reference standard curve

### CSF Total Tau Assay Performance

To benchmark the CSF Total Tau assay compared to previously published methods using PCA precipitation, we performed a serial dilution of Tau 441 in artificial CSF (0.5% serum in PBS), an artificial matrix used in similar protocols for quantification of tau peptides against a reference standard [17]. In three independent analytical runs, we defined the linear range for each surrogate peptide where mean bias and CV were ≤ 20% (Table 5). The dynamic range for each surrogate peptide was 2 – 3 orders of magnitude in artificial CSF surrogate matrix. The lower limits of quantification for each peptide were in the range of hundreds of pg/mL to single ng/mL, as previously reported [18]. These data suggest that our CSF Total Tau Assay shows a linear response over the range of endogenous CSF Tau surrogate peptide concentrations.

We next spiked in known concentrations of recombinant tau 441 to non-disease human CSF to test for matrix effects compared to our reference standard. Endogenous CSF Tau surrogate peptide concentrations within the defined linear range from the reference standard curve ranged from 797 pg/mL (396-406) to 12,358 pg/mL (181-190), and one calculated peptide concentration (243-254) was detected below the lower limit of quantification on the reference standard curve. Moreover, 6 of the 7 peptides met analytical acceptance criteria of CV ≤ 30% (Table 5). Although we defined linear ranges for multiple peptides < 1,000 pg/mL in surrogate CSF using recombinant tau, the mean bias for non-disease human CSF plus 1,000 pg/mL Tau 441 did not meet analytical acceptance criteria of ≤ 30% for all peptides analyzed. However, all peptides had a relative error ≤ 30% for the other two spike-in conditions (+ 4,000 pg/mL, + 16,000 pg/mL). Taken together, these data confirm the ability to measure recombinant tau 441 in artificial CSF surrogate matrix at sensitivities similar to previous reports [18]. However, the inability to accurately quantify 1,000 pg/mL of recombinant tau in CSF suggests a lack of assay parallelism between surrogate and CSF matrix.

### Linearity and Parallelism in Alzheimer’s Disease CSF

We next tested for parallelism of recombinant Tau 441 in artificial CSF with endogenous tau in Alzheimer’s Disease CSF for 6 out of the 7 peptides that were detected in the two AD CSF samples (Figure 2). Peptide 354-369 was excluded from this analysis because a peak was not detected in the AD CSF samples. 4 peptides demonstrated a linear concentration-response of calculated concentration in both AD CSF sample dilution series (25-44, 181-190, 212-221, 243-254), as evidenced by reported R^2^ ≥ 0.85 for each dilution curve (Figures 2A-D). However, none of the 6 peptides assayed demonstrated parallelism of endogenous tau with the recombinant tau 441 in artificial CSF (Figure 2A-F). These data suggest that this assay can successfully determine abundance changes of endogenous tau surrogate peptides 25-44, 181-190, 212-221 and 243-254, but that the reference standard curve does not accurately quantify the true tau concentration in Alzheimer’s Disease CSF. According to the Lee *et al*. publication on biomarker method validation, this would meet the criteria of a quasi-quantitative method [21].

**Figure 2.**
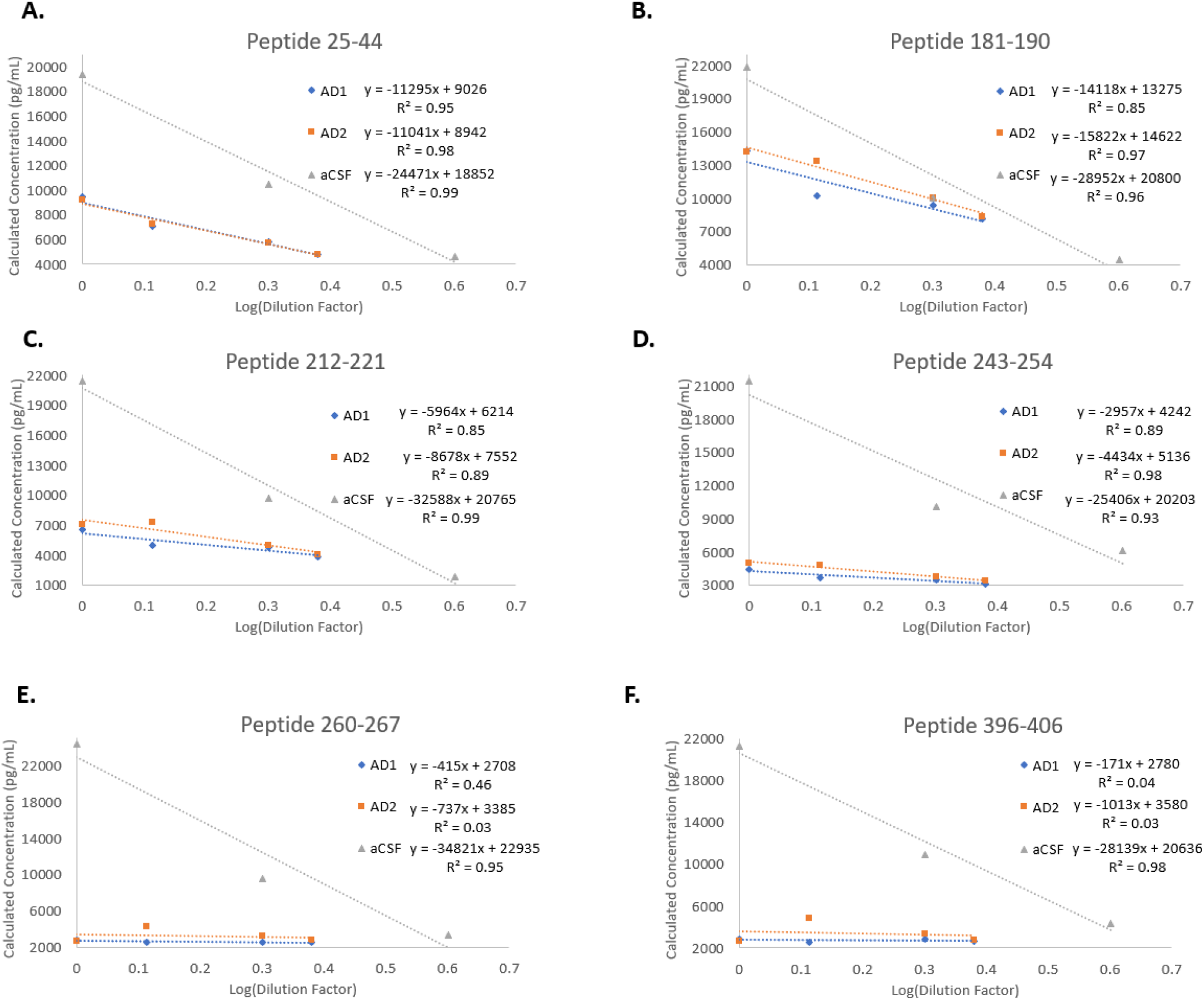
Test of dilutional linearity and parallelism between endogenous tau protein in Alzheimer’s Disease CSF and recombinant tau 441 in artificial CSF. A linear regression analysis of calculated tau concentration versus dilution factor was performed on AD CSF and artificial CSF matrices and compared for six tau surrogate peptides. Peptide 354-369 was not identified in either undiluted AD CSF sample, so it was excluded from analysis. A. Surrogate peptide 25-44 demonstrates dilutional linearity in all matrices, but AD CSF and reference standard curve are not parallel. B. Surrogate peptide 181-190 demonstrates dilutional linearity in all matrices, but AD CSF and reference standard curve are not parallel. C. Surrogate peptide 212-221 demonstrates dilutional linearity in all matrices, but AD CSF and reference standard curve are not parallel. D. Surrogate peptide 243-254 demonstrates dilutional linearity in all matrices, but AD CSF and reference standard curve are not parallel. E. Surrogate peptide 260-267 does not show dilutional linearity in AD CSF samples. F. Surrogate peptide 396-406 does not show dilutional linearity in AD CSF samples.

### Fit-for-purpose Biomarker Assay Determination

We synthesized the results from the two parallelism experiments: one in human CSF with recombinant tau spiked-in (Table 5), and one in AD CSF monitoring endogenous tau (Figure 2) to determine the biomarker categorization of each surrogate peptide analysis in our multiplexed LC-MS assay. Our results suggest that LC-MS analysis of peptides 25-44, 181-190, 212-221, and 243-254 can be categorized as quasi quantitative biomarker assays. For quasi quantitative analysis, we demonstrated a linear range to quantify differences in surrogate peptide abundance in AD CSF (Figure 2), but the reference standard does not accurately quantify the absolute concentration of tau based on these surrogate peptides (Figure 2A-D). Surrogate peptides 260-267 and 396-406 can be categorized as qualitative assays, where the assay can be used to determine the presence or absence of tau species containing these amino acid sequences above the linear range described by the reference standard curve but not to compare abundance changes between biological states, as demonstrated by a lack of signal proportionality in the AD CSF parallelism experiments (Figure 2E-F).

### CSF Total Tau in Alzheimer’s Disease CSF

We next assayed CSF Total Tau peptide concentrations in 11 Alzheimer’s Disease CSF samples across two independent analytical runs (n = 4 replicates) to understand the precision of our LC-MS assay in diseased CSF (Table 6). We observed signal detection of peptides 25-44, 181-190, and 212-221 in all 11 samples analyzed, and a lower percentage of 243-254, 260-267 and 396-406 across the 11 samples. CSF tau peptides defined as quasi-quantitative biomarker assays (25-44, 181-190, 212-221, and 243-254) varied in their performance, as measured by sample variance. When signal detection of peptide 243-254 was observed in a sample, CV values were < 20%. For peptide 25-44 and 181-190, CV < 33% for 10/11 samples. For peptide 212-221, CV < 30% for 8/11 peptides, and CV was above 45% for other 3 samples. In sum, these data demonstrate the differential analytical capabilities of each surrogate peptide by LC-MS/MS in Alzheimer’s Disease CSF.

**Table 6.** CSF Total Tau Analysis in Alzheimer’s Disease CSF. Reported values represent the inter-assay mean calculated concentration and variance across the two analytical runs (n = 4 replicates). + peak detected for qualitative analysis, - no peak detection in sample.

| AD CSF<br>Sample<br>Number: | 1 | 2 | 3 | 4 | 5 | 6 | 7 | 8 | 9 | 10 | 11 | Total |
| --- | --- | --- | --- | --- | --- | --- | --- | --- | --- | --- | --- | --- |
| Surrogate<br>Peptide | Concentration<br>(pg/mL)<br>(% CV) | Concentration<br>(pg/mL)<br>(% CV) | Concentration<br>(pg/mL)<br>(%CV) | Concentration<br>(pg/mL)<br>(% CV) | Concentration<br>(pg/mL)<br>(% CV) | Concentration<br>(pg/mL)<br>(% CV) | Concentration<br>(pg/mL)<br>(% CV) | Concentration<br>(pg/mL)<br>(% CV) | Concentration<br>(pg/mL)<br>(% CV) | Concentration<br>(pg/mL)<br>(% CV) | Concentration<br>(pg/mL)<br>(% CV) | % Samples<br>Peptide<br>Detected<br>(n) |
| 25-44 | 9,680 (21) | 8,830 (10) | 2,859 (11) | 1,873 (13) | 3,984 (4) | 2,264 (18) | 3,973 (30) | 3,906 (16) | 2,238 (9) | 2239 (31) | 2627 (43) | 100 (11) |
| 181-190 | 12,469 (10) | 13,225 (9) | 4,506 (2) | 3,647 (2) | 5,128 (5) | 3,616 (16) | 8,104 (27) | 5,917 (4) | 3,800 (15) | 3780 (15) | 4682 (41) | 100 (11) |
| 212-221 | 6,742 (17) | 7,407 (7) | 1,501 (25) | 1,587 (24) | 2,618 (17) | 1,397 (46) | 2,645 (25) | 3,069 (7) | 1,539 (17) | 1,217 (57) | 1,720 (52) | 100 (11) |
| 243-254 | 2,378 (12) | 3,090 (10) | - | - | 1,196 (6) | - | - | 1,385 (14) | - | - | - | 36 (4) |
| 260-267 | + | + | - | - | - | - | - | - | - | - | - | 18 (2) |
| 354-369 | - | - | - | - | - | - | - | - | - | - | - | 0 (0) |
| 396-406 | + | + | - | - | - | - | - | - | - | - | - | 18 (2) |

### Therapeutic Antibody Interference

To assess the utility of our CSF Total Tau assay in the presence of therapeutic antibody, we performed an *ex vivo* incubation of non-disease human CSF with varying concentrations of ABBV-8E12 to model the sample conditions of a CSF sample collected in an anti-tau monoclonal antibody clinical trial (Table 7). For all quasi quantitative peptides assayed, the mean bias between the endogenous CSF Tau samples with and without ABBV-8E12 was ≤ 10%. Therefore, for the peptides categorized as quasi quantitative biomarker assays (25-44, 181-190,212-221, and 243-254), these data suggest that the CSF Total Tau Assay can reliably monitor these N-Terminus and Mid-Domain peptides without interference by therapeutic monoclonal antibodies.

**Table 7.** CSF Total Tau interference test in the presence of anti-tau monoclonal antibody ABBV-8E12. Reported values represent the mean calculated concentration and variance across one analytical run (n = 3 replicates).

| Surrogate Peptide | Calculated CSF Tau Concentration (pg/mL) |  |  |  |  |  |  |  |
| --- | --- | --- | --- | --- | --- | --- | --- | --- |
|  | + 0 ug/mL ABBV-8E12 | CV (%) | + 2.5 ug/mL ABBV-8E12 | CV (%) | Mean Bias (%) | +5.0 ug/mL ABBV-8E12 | CV (%) | Mean Bias (%) |
| 25-44 | 8,248 | 3 | 7,699 | 6 | 7 | 7,974 | 4 | 3 |
| 181-190 | 12,856 | 1 | 12,561 | 2 | 2 | 12,239 | 2 | 5 |
| 212-221 | 5,109 | 8 | 4,931 | 2 | 3 | 5,004 | 5 | 2 |
| 243-254 | 2,686 | 11 | 2,480 | 2 | 8 | 2,571 | 2 | 4 |

## Discussion

In the setting of clinical trial bioanalysis, increase in sample throughput is a crucial consideration. Additionally, modified parameters such as the 1% PCA precipitation directly to enzymatic digestion makes the method more suitable for existing bioanalytical workflows such as antibody pharmacokinetics measures. In the pharmaceutical industry, where operational efficiency is a key criteria to optimize in method development, our workflow provides significant advantages in sample throughput, as our sample preparation protocol takes 48 hours to completion, contrary to Barthelemy et al., which uses a 72 hour sample preparation protocol[17].

The method described here quantifies seven peptides representing multiple functional domains of tau protein, from N-to C-terminus in human cerebrospinal fluid (CSF). Our method demonstrated similar measures of sensitivity in surrogate matrix with recombinant protein standards as previously published protocols that aimed to use a CSF Total Tau assay in the pharmaceutical industry setting [18], with the inclusion of an N-Terminus peptide that is crucial for assaying target binding of N-terminal targeting monoclonal antibodies. Similar to Bros et al.[18], we observed peak interference in attempts to identify peptide 25-44 using a triple quadrupole mass analyzer (data not shown). The ability to detect this peptide was afforded due to the selection of a high resolution mass spectrometer [23], supporting the use of orbitrap mass analyzers for biomarker measurements in the pharmaceutical bioanalytical space. Although convention is to use a triple quadrupole mass analyzer, our data establish further strength to the argument that parallel reaction monitoring methods are robust and reproducible for measuring endogenous protein-based biomarkers in pharmaceutical bioanalysis.

Our data corroborate previous studies that utilize perchloric acid as an immunoenrichment-free sample preparation method, with differential detection capabilities of N-terminus and mid domain peptides detected extracellularly compared to peptides at the C terminus [23-25]. However, to the best of our knowledge, no studies directly tested parallelism of endogenous tau protein in CSF to validate these calculated concentrations as accurate measures of AD CSF tau surrogate peptide abundances. Barthelemy et al. demonstrated matrix effects of lowered signal intensity of recombinant tau 441 in human CSF compared to artificial CSF, and used this difference in signal intensity as a scaling factor to determine the LLOQ of endogenous tau peptides in human CSF [17]. However, this approach did not directly test the concentration-response curve of endogenous tau. Our demonstration of the lack of parallelism between endogenous tau in AD CSF compared to recombinant tau in surrogate matrix suggest that differences in tau surrogate peptide abundances may only be compared within one surrogate peptide between samples, as opposed to comparing the abundance of N-Terminus peptides compared to Mid-domain and C-Terminus peptides. Despite a lack of parallelism, we were able to establish dilutional linearity in AD CSF for 4 surrogate peptides, and our ability to detect a range of quasi quantitative concentration values in AD CSF samples suggests that our assay is sufficient for generating data on changes in endogenous tau protein concentration using surrogate peptides that add proteoform resolution beyond single epitope LBA-based assays. The linear range defined in our assay is limited by the AD CSF pool used as a diluent that had a baseline level of tau protein itself. Therefore, future investigations should aim to use AD CSF depleted of tau protein, or other surrogate matrices that can determine the lower linear range for the quasi-quantitative peptides.

Despite its advantages, our assay does exhibit some limitations, including a decreased sensitivity and limited panel of surrogate peptides compared to the panel Barthelemy et al. have analyzed using similar methods[17,23]. Additionally, nanoflow LC and other enrichment strategies achieve lower LLOQ than what we showed here with higher flow UHPLC[9,26]. Our assay requires 500 µL of CSF, a precious and limited biofluid from individuals in clinical trials. In this regard, we do not believe the CSF Tau Assay defined in this report should be used as a diagnostic biomarker, where established assays are in place with superior sensitivity by both LBA and LC-MS. However, tau concentration will likely be increased in anti-Tau-dosed subjects, as therapeutic monoclonal antibodies are expected to increase the concentration of tau in biofluids due to increased stability of monoclonal antibodies in circulation compared to tau protein itself [27]. The CV values defined for each quasi-quantitative CSF tau peptide assay in AD CSF suggests that our assay will reliably detect ≥ 1.5 fold increases of tau peptide concentrations in dosed anti-tau subjects relative to baseline values. Therefore, we believe this assay should be used for pharmacodynamics biomarker assessment to better understand exposure-response relationships, as it provides proteoform information beyond current LBA-based assays.

Many current efforts use ligand binding assays (LBA) with a combination of two epitopes analyzed by capture and detection antibodies [7]. Although these assays achieve superior sensitivity for relative quantitative assays compared to LC-MS, any analysis using these assays will ultimately be limited to the tau species containing these two antibody epitopes. In the complex disease biology of primary tauopathies, where tau protein presents as multiple proteoforms in biofluid samples, it is important to develop multiplexed assays that explore tau pharmacodynamics to determine the relevant pathogenic species to target in human disease. Moreover, careful selection of capture and detection antibodies must be considered in relationship to the therapeutic antibody, creating a complicated method development strategy that requires custom assay development for screening monoclonal antibodies targeting different regions of a protein. Our method provides a simple solution to overcome these time-consuming complexities, as we detected no interference for our LC-MS assay to calculate CSF total tau concentrations in the presence of anti-Tau antibody (Table 7).

Further work should also aim to increase the proteoform resolution of this assay, particularly for phosphorylated [28-30] and differentially cleaved [29,31-33] tau proteoforms that are emerging as diagnostic biomarkers for Alzheimer’s Disease. Incorporation of phosphorylated and semi-tryptic peptides into an LC-MS approach would be invaluable for testing hypotheses that these phosphorylation sites have utility as both diagnostic and pharmacodynamic biomarkers. However, as is the case with all bottom-up surrogate peptide LC-MS analyses, these assays are limited in their ability to draw conclusions on the exact proteoform of the analytes measured in the assay[34]. Despite these limitations, correlations of the changes in peptide abundances in the presence of therapeutic intervention may provide insight into the key posttranslational modifications/proteoform features that define effective disease modification strategies.

## Conclusions

The CSF Total Tau LC-MS method described here successfully achieved the fit-for-purpose biomarker specifications needed for a total tau bioanalysis in the presence of therapeutic monoclonal antibodies. In sum, we believe that LC-MS based assays to quantify extracellular tau peptides hold great promise as a translatable bioanalytical tool for future target binding studies in preclinical models of tauopathy and human clinical trials.

## Data Availability

All relevant data are presented in the manuscript.

## Acknowledgements

We thank Klaus Magin at AbbVie for contributions in optimizing sample preparation conditions for the final assay workflow. We also thank Dr. Mario Richter and Dr. Gary Jenkins at AbbVie for their insight and discussion around these data.

## Ethical Conduct of Research Statement

AbbVie is committed to the internationally-accepted standard of the 3Rs (Reduction, Refinement, Replacement) and adhering to the highest standards of animal welfare in the company’s research and development programs. Animal studies were approved by AbbVie’s [Institutional Animal Care and Use Committee or Ethics Committee]. Animal studies were conducted in an AAALAC accredited program where veterinary care and oversight was provided to ensure appropriate animal care.

## References

1. Wang Y, Mandelkow E. Tau in physiology and pathology. Nature Reviews Neuroscience, 17(1), 22–35 (2016).

2. Gordon BA, Blazey TM, Christensen J et al. Tau PET in autosomal dominant Alzheimer’s disease: relationship with cognition, dementia and other biomarkers. Brain : a journal of neurology, 142(4), 1063–1076 (2019).

3. La Joie R, Visani AV, Baker SL et al. Prospective longitudinal atrophy in Alzheimer’s disease correlates with the intensity and topography of baseline tau-PET. Science translational medicine, 12(524) (2020).

4. Braak H, Braak E. Neuropathological stageing of Alzheimer-related changes. Acta neuropathologica, 82(4), 239–259 (1991).

5. Mirbaha H, Chen D, Morazova OA et al. Inert and seed-competent tau monomers suggest structural origins of aggregation. eLife, 7 (2018).

6. Rauch JN, Luna G, Guzman E et al. LRP1 is a master regulator of tau uptake and spread. Nature, 580(7803), 381–385 (2020).

7. Boxer AL, Qureshi I, Ahlijanian M et al. Safety of the tau-directed monoclonal antibody BIIB092 in progressive supranuclear palsy: a randomised, placebo-controlled, multiple ascending dose phase 1b trial. The Lancet Neurology, 18(6), 549–558 (2019).

8. West T, Hu Y, Verghese PB et al. Preclinical and Clinical Development of ABBV-8E12, a Humanized Anti-Tau Antibody, for Treatment of Alzheimer’s Disease and Other Tauopathies. The journal of prevention of Alzheimer’s disease, 4(4), 236–241 (2017).

9. Sato C, Barthelemy NR, Mawuenyega KG et al. Tau Kinetics in Neurons and the Human Central Nervous System. Neuron, 97(6), 1284-1298.e1287 (2018).

10. Goedert M, Spillantini MG, Jakes R, Rutherford D, Crowther RA. Multiple isoforms of human microtubule-associated protein tau: sequences and localization in neurofibrillary tangles of Alzheimer’s disease. Neuron, 3(4), 519–526 (1989).

11. Fitzpatrick AWP, Falcon B, He S et al. Cryo-EM structures of tau filaments from Alzheimer’s disease. Nature, 547(7662), 185–190 (2017).

12. Arakhamia T, Lee CE, Carlomagno Y et al. Posttranslational Modifications Mediate the Structural Diversity of Tauopathy Strains. Cell, (2020).

13. Blennow K, Hampel H, Weiner M, Zetterberg H. Cerebrospinal fluid and plasma biomarkers in Alzheimer disease. Nature reviews. Neurology, 6(3), 131–144 (2010).

14. Khoonsari PE, Shevchenko G, Herman S et al. Improved Differential Diagnosis of Alzheimer’s Disease by Integrating ELISA and Mass Spectrometry-Based Cerebrospinal Fluid Biomarkers. Journal of Alzheimer’s disease : JAD, 67(2), 639–651 (2019).

15. Johnson GV, Seubert P, Cox TM, Motter R, Brown JP, Galasko D. The tau protein in human cerebrospinal fluid in Alzheimer’s disease consists of proteolytically derived fragments. Journal of neurochemistry, 68(1), 430–433 (1997).

16. Meredith JE Jr.,, Sankaranarayanan S, Guss V et al. Characterization of novel CSF Tau and ptau biomarkers for Alzheimer’s disease. PloS one, 8(10), e76523 (2013).

17. Barthelemy NR, Fenaille F, Hirtz C et al. Tau Protein Quantification in Human Cerebrospinal Fluid by Targeted Mass Spectrometry at High Sequence Coverage Provides Insights into Its Primary Structure Heterogeneity. Journal of proteome research, 15(2), 667–676 (2016).

18. Bros P, Vialaret J, Barthelemy N et al. Antibody-free quantification of seven tau peptides in human CSF using targeted mass spectrometry. Frontiers in neuroscience, 9, 302 (2015).

19. Barghorn S, Biernat J, Mandelkow E. Purification of recombinant tau protein and preparation of Alzheimer-paired helical filaments in vitro. Methods Mol Biol, 299, 35–51 (2005).

20. Lindwall G, Cole RD. The purification of tau protein and the occurrence of two phosphorylation states of tau in brain. Journal of Biological Chemistry, 259(19), 12241–12245 (1984).

21. Lee JW, Devanarayan V, Barrett YC et al. Fit-for-Purpose Method Development and Validation for Successful Biomarker Measurement. Pharmaceutical Research, 23(2), 312–328 (2006).

22. Ivanovova N, Handzusova M, Hanes J, Kontsekova E, Novak M. High-yield purification of fetal tau preserving its structure and phosphorylation pattern. Journal of immunological methods, 339(1), 17–22 (2008).

23. Barthelemy NR, Gabelle A, Hirtz C et al. Differential Mass Spectrometry Profiles of Tau Protein in the Cerebrospinal Fluid of Patients with Alzheimer’s Disease, Progressive Supranuclear Palsy, and Dementia with Lewy Bodies. Journal of Alzheimer’s disease : JAD, 51(4), 1033–1043 (2016).

24. Portelius E, Hansson SF, Tran AJ et al. Characterization of Tau in Cerebrospinal Fluid Using Mass Spectrometry. Journal of proteome research, 7(5), 2114–2120 (2008).

25. McAvoy T, Lassman ME, Spellman DS et al. Quantification of tau in cerebrospinal fluid by immunoaffinity enrichment and tandem mass spectrometry. Clinical chemistry, 60(4), 683–689 (2014).

26. Zhou M, Duong DM, Johnson ECB et al. Mass Spectrometry-Based Quantification of Tau in Human Cerebrospinal Fluid Using a Complementary Tryptic Peptide Standard. Journal of proteome research, (2019).

27. Yanamandra K, Patel TK, Jiang H et al. Anti-tau antibody administration increases plasma tau in transgenic mice and patients with tauopathy. Science translational medicine, 9(386) (2017).

28. Barthélemy NR, Bateman RJ, Hirtz C et al. Cerebrospinal fluid phospho-tau T217 outperforms T181 as a biomarker for the differential diagnosis of Alzheimer’s disease and PET amyloid-positive patient identification. Alzheimer’s research & therapy, 12(1), 26 (2020).

29. Blennow K, Chen C, Cicognola C et al. Cerebrospinal fluid tau fragment correlates with tau PET: a candidate biomarker for tangle pathology. Brain : a journal of neurology, 143(2), 650–660 (2019).

30. Janelidze S, Mattsson N, Palmqvist S et al. Plasma P-tau181 in Alzheimer’s disease: relationship to other biomarkers, differential diagnosis, neuropathology and longitudinal progression to Alzheimer’s dementia. Nature medicine, 26(3), 379–386 (2020).

31. Schlegel K, Awwad K, Heym RG et al. N368-Tau fragments generated by legumain are detected only in trace amount in the insoluble Tau aggregates isolated from AD brain. Acta neuropathologica communications, 7(1), 177 (2019).

32. Zhang Z, Song M, Liu X et al. Cleavage of tau by asparagine endopeptidase mediates the neurofibrillary pathology in Alzheimer’s disease. Nature medicine, 20(11), 1254–1262 (2014).

33. Chen HH, Liu P, Auger P et al. Calpain-mediated tau fragmentation is altered in Alzheimer’s disease progression. Sci Rep, 8(1), 16725 (2018).

34. Savaryn JP, Catherman AD, Thomas PM, Abecassis MM, Kelleher NL. The emergence of top-down proteomics in clinical research. Genome medicine, 5(6), 53 (2013).

35. Yamada K, Cirrito JR, Stewart FR et al. In vivo microdialysis reveals age-dependent decrease of brain interstitial fluid tau levels in P301S human tau transgenic mice. The Journal of neuroscience : the official journal of the Society for Neuroscience, 31(37), 13110–13117 (2011).

